# Interpretable Hazard Models Reveal Strong Metastasis Dependence and Feature Interaction Effects in Predicting Cancer Patient Readmission Risk

**DOI:** 10.1101/2025.06.23.25330159

**Authors:** Sibei Liu, Xintao Li, Zhijian Hu

**Affiliations:** Miami Herbert Business School, University of Miami, Coral Gables, USA; College of Computing, Georgia Institute of Technology, Atlanta, USA; Department of Biophysics, University of Michigan, Ann Arbor, USA

## Abstract

Predicting hospital readmission in cancer patients-particularly those with metastatic disease-remains a significant clinical challenge. While metastasis is known to affect prognosis, its role in shaping the relationship between standard clinical features (e.g., lab tests) and readmission risk remains poorly understood. In this study, we leverage the MIMIC-IV database to stratify patients by metastatic burden, defined by the number of metastatic sites, and analyze survival dynamics using a two-step neural-symbolic distillation framework. First, assuming the proportional hazards structure, we train a neural network to learn both the baseline hazard *h*_0_(*t*) and the log-partial hazard *f* (*x*), capturing complex, noisy survival patterns. We then distill the learned model into interpretable symbolic functions using Kolmogorov-Arnold Networks (KANs). For *h*_0_(*t*), we observe metastasis-specific linear or quadratic trends, with significantly steeper early-time hazards in patients with three or more metastatic sites-highlighting the need for enhanced post-discharge monitoring. To simplify the symbolization of *f* (*x*), we introduce a prestructuring step using B -spline expansions with both univariate and interaction terms. This reduces symbolic complexity and reveals that interaction effects dominate, with larger coefficients than univariate terms in most subgroups. The resulting symbolic expressions for *f* (*x*) vary substantially across metastasis strata, indicating that the influence of lab-derived features on readmission is metastasis-dependent. Our framework offers interpretable, metastasis-stratified survival models and provides a foundation for deeper exploration of lab-metastasis interactions in personalized cancer care.

## Introduction

Hospital readmissions remain a critical issue for healthcare systems worldwide, particularly among cancer patients whose complex medical needs often result in frequent returns to care. Previous studies reveal high readmission rates within the first months following discharge; nearly 20% of Medicare beneficiaries, for example, are readmitted within 30 days, with rates reaching over 50% within the first year [1]. Cancer patients, especially those with advanced or metastatic disease, face even greater readmission risks, placing additional pressure on healthcare resources in both the United States and Canada [2–5]. The economic impact of these readmissions is substantial. In the United States, unplanned readmissions have accounted for over 40 billion US dollars in hospital expenses, a significant portion of which is linked to patients with chronic or progressive diseases, such as cancer [6, 7]. The need to manage both the cost and complexity of cancer patient readmissions highlights the importance of accurate prediction models that can anticipate readmission timing and tailor post-discharge care. In the US, the Centers for Medicare and Medicaid Services (CMS) established penalties for hospitals with high 30-day readmission rates by reducing the payment for readmitted patients [8].

Laboratory biomarkers play a critical role in assessing cancer patients’ condition and have shown significant predictive power for hospital readmissions. Abnormal blood test results often reflect underlying complications or poor physiological reserve, which can foreshadow an early return to the hospital. Recent research demonstrates that incorporating even routine lab parameters (e.g. hemoglobin, white blood cell counts, platelets, etc.) into risk models markedly improves predictive accuracy across multiple cancer types [9]. For instance, older oncology patients with at least two abnormal lab values at discharge (such as low hemoglobin, hypoalbuminemia, or electrolyte imbalances) were found to have 3-fold higher odds of 30-day readmission compared to those with one or no abnormalities [10]. Likewise, a dedicated cancer readmission risk score identified anemia (hemoglobin < 10*g/dL*) and leukocytosis (WBC > 11× 10^9^*/L*) as independent predictors of unplanned readmission [11], underscoring how deviations in common lab measures signal elevated risk. However, most such models consider each lab metric in isolation – typically flagging individual derangements against fixed thresholds and do not explicitly evaluate interaction effects between different lab features. In other words, current approaches tend to sum or count abnormal results for a general risk estimate [10], but rarely examine whether specific combinations of lab abnormalities, or even seemingly normal lab results, synergistically exacerbate readmission risk beyond their individual contributions. This gap suggests that while lab tests are good indicators on their own, there is still a need to explore how concurrent lab derangements, or even normal lab features, might interact to better stratify readmission risk in cancer patients.

Traditional survival models, such as the Cox proportional hazards (CPH) model, have been widely used to predict patient readmission risk and survival outcomes in healthcare [12–15]. However, the classic Cox model assumes that the log-partial hazard is a linear function of the covariates [16]. In practice, it may fail to capture complex non-linear or interactive effects of patient variables on the risk. To better model complex relationships in the data, researchers have increasingly turned to machine learning (ML) and deep learning approaches for survival analysis and readmission risk prediction. Methods such as random survival forests (an extension of random forests for time-to-event data) and recurrent neural networks like LSTMs have been applied to capture non-linear interactions and time-dependent features that a linear Cox model might miss [17–22]. Several modern survival models also try to combine the Cox framework with the flexibility of deep learning. For example, DeepSurv extends the Cox PH model by using a deep feed-forward neural network to learn a non-linear log-partial hazard function instead of a simple linear combination of covariates [23]. Another model, DeepHit, takes a different approach by learning the entire distribution of survival times with a deep neural network, without requiring the proportional hazards assumption [24]. These approaches often achieve improved predictive accuracy in high-dimensional settings by relaxing the linear assumption and learning nonlinear effects. However, a major drawback is interpretability: these models are typically treated as “black boxes,” making it difficult to understand how they arrive at a given risk prediction. Unlike the straightforward coefficients and hazard ratios from a Cox model, complex ensemble or neural network models do not readily explain the contribution of each input feature to the outcome. This lack of transparency can hinder clinical trust and adoption of such models, especially in high-stakes medical decision making. To address this gap, researchers have developed a variety of strategies to interpret or explain survival risk models. Model-agnostic explanation techniques such as LIME and SHAP (SHapley Additive exPlanations) have been applied to survival predictions to quantify the influence of each feature on an individual patient’s risk [18, 25, 26]. However, these post-hoc interpretability tools still do not capture the full relationship learned by complex models. The explanations are often local (explaining one prediction at a time) or feature-centric, and may not neatly describe higher-order interactions or the way multiple features jointly influence risk over time [27].

Although readmission risks for cancer patients across different cancer types have been extensively studied [3, 28, 29], the joint influence of metastatic burden—measured by the number of metastatic sites—and standard laboratory test results on readmission risk remains largely unexplored. In this work, we investigate how metastasis–lab interactions and lab–lab interactions shape readmission risk by applying a novel neural-symbolic distillation framework that yields interpretable functional relationships. Using the MIMIC-IV database, we first perform exploratory correlation analysis between readmission days, metastatic burden, and lab-derived features. As expected, neither the metastatic burden nor individual lab features show strong linear correlations with readmission time—suggesting that single features alone are insufficient for reliable prediction. This highlights the likely importance of higher-order interactions. To uncover these effects, we develop a two-step neural-symbolic distillation framework. First, under the Proportional Hazards Assumption, we train a neural network to model both the baseline hazard *h*_0_(*t*; *θ*) and the log-partial hazard *f* (*x*; *θ*). We then distill the learned model into compact, interpretable symbolic functions using Kolmogorov–Arnold Networks (KANs). For *h*_0_(*t*), we observe metastasis-specific linear or quadratic trends, with notably steeper slopes for patients with ≥3 metastatic sites—highlighting a critical post-discharge period requiring close monitoring. To aid symbolic recovery of *f* (*x*), we introduce a prestructuring step using B-spline expansions that include both univariate terms and pairwise interaction terms—akin to a structured ansatz in physics. This reduces the complexity of the symbolic learning task and consistently reveals that interaction terms carry substantially larger coefficients than univariate terms, underscoring the importance of nonlinear, multivariate effects. The resulting symbolic expressions for *f* (*x*) differ markedly across metastasis strata, indicating that lab-feature influence on readmission is strongly metastasis-dependent. Importantly, all models are trained with minimal hyperparameter tuning and standard network configurations, suggesting robustness and generalizability of the proposed method.

## Highlights

- Recent studies have shown that traditional proportional hazards models often violate the proportional hazards assumption in real-world datasets, underscoring the need for stratified or subgroup-specific analyses.
- Cox Proportional Hazards (CoxPH) models offer high interpretability but limited capacity to capture nonlinear effects. In contrast, neural network-based approaches such as DeepSurv can model complex, nonlinear survival patterns but lack transparency. This study integrates these paradigms through a neural-symbolic distillation framework to achieve both interpretability and predictive performance in cancer readmission risk modeling.
- Our findings highlight the prognostic importance of metastatic burden and feature–feature interactions. These interactions may inspire the development of new interpretable prognostic scores based on lab-derived feature combinations—analogous to established clinical indices such as the Neutrophil-to-Lymphocyte Ratio (NLR) or Pan-Immune-Inflammation Value (PIV).

## Methods

### Data Source and Preprocessing

The dataset used in this study was derived from the MIMIC-IV database [30], which contains de-identified health records for thousands of hospital admissions, including demographic information, laboratory test results, and ICD-9/10 diagnosis codes.

### Data Cleaning

- We began by extracting 37,623 admissions corresponding to 18,125 unique patients diagnosed with one or more of 13 common cancer types. These include breast, lung, colorectal, pleura, mediastinum, respiratory, urinary, bladder, skin, nervous system, bone, female genital, and male genital cancers (denoted in the dataset as `Breast Cancer`, `Lung Cancer`, `Colorectal Cancer`, etc.). Cancer types were determined based on ICD codes following the National Cancer Institute’s case-finding rules. Each cancer type variable is encoded as 1 for primary and 2 for metastatic diagnoses per admission.
- Two variables were available for readmission status: a binary indicator of whether the patient was ever readmitted, and a numerical variable (`y readmission day`) capturing the number of days between admissions. Patients who were not readmitted during the observation window were treated as right-censored in the survival analysis. We used anchor age and the maximum discharge time in MIMIC-IV to estimate the maximum observed follow-up time for these patients. Their `y readmission day`values were set accordingly.
- To ensure clinically meaningful readmission timelines, we excluded admissions where the time to readmission exceeded 1,095 days (3 years). Features with more than 25% missing values were removed to reduce the influence of data sparsity and noise. For remaining features with missing values, we applied median imputation. This preprocessing resulted in the selection of 11 key lab-derived features: `first blood pressure`, `first height`, `platelet`, `white blood cell`, `hemoglobin`, `glucose`, `bicarbonate`, `creatinine`, `chloride`, `potassium`, and `sodium`.
- To mitigate scale differences and facilitate training, we standardized all lab-derived features using the `StandardScaler`from `scikit-learn`, transforming each feature to have zero mean and unit variance. This transformation preserves interpretability, as values can be easily reverted to their original scales through inverse transformation when needed.
- To investigate the relationship between metastatic burden and readmission dynamics, we stratified patients into four groups based on the number of metastatic sites:
  ‐ Group 1: No metastases (*n* = 0)
  ‐ Group 2: One metastatic site (*n* = 1)
  ‐ Group 3: Two metastatic sites (*n* = 2)
  ‐ Group 4: Three or more metastatic sites (*n ≥* 3)

This stratification was applied regardless of cancer type to enable analysis of how disease severity influences readmission risks and survival trajectories.

After data cleaning, we derived a standardized dataset consisting of 11 lab-derived features and four metastastic burden subgroups. The dataset includes a total of 18,704 uncensored admission entries and 2,432 right-censored entries. Since the number of uncensored entries far exceeds that of censored cases, and to avoid complications due to class imbalance, we focus exclusively on the uncensored subset for simplicity. The final dataset thus comprises 18,704 admissions, distributed across metastastic categories as follows: 7,102 entries for patients with no metastasis (*n* = 0), 6,441 entries with one metastatic tumor (*n* = 1), 3,420 entries with two metastatic tumors (*n* = 2), and 1,741 entries with three or more metastatic tumors (*n* ≥ 3). In the following text, we use the terms *metastastic burden, metastastic tumor*, and *metastastic site* interchangeably to refer to different levels of metastatic status.

### Symbolic Survival Learning from Metastasis-Lab Data

In this study, we present a novel, interpretable framework for predicting hospital readmission risks in cancer patients, stratified by metastatic burden. Our workflow (Figure 1B) combines traditional survival analysis with deep neural modeling and symbolic regression techniques to extract interpretable patterns from noisy clinical data. We begin with preprocessed data from the MIMIC-IV database (n = 18704), consisting of cancer patient records annotated by metastasis status and a panel of standardized laboratory test features selected during the preprocessing.

**Figure 1.**
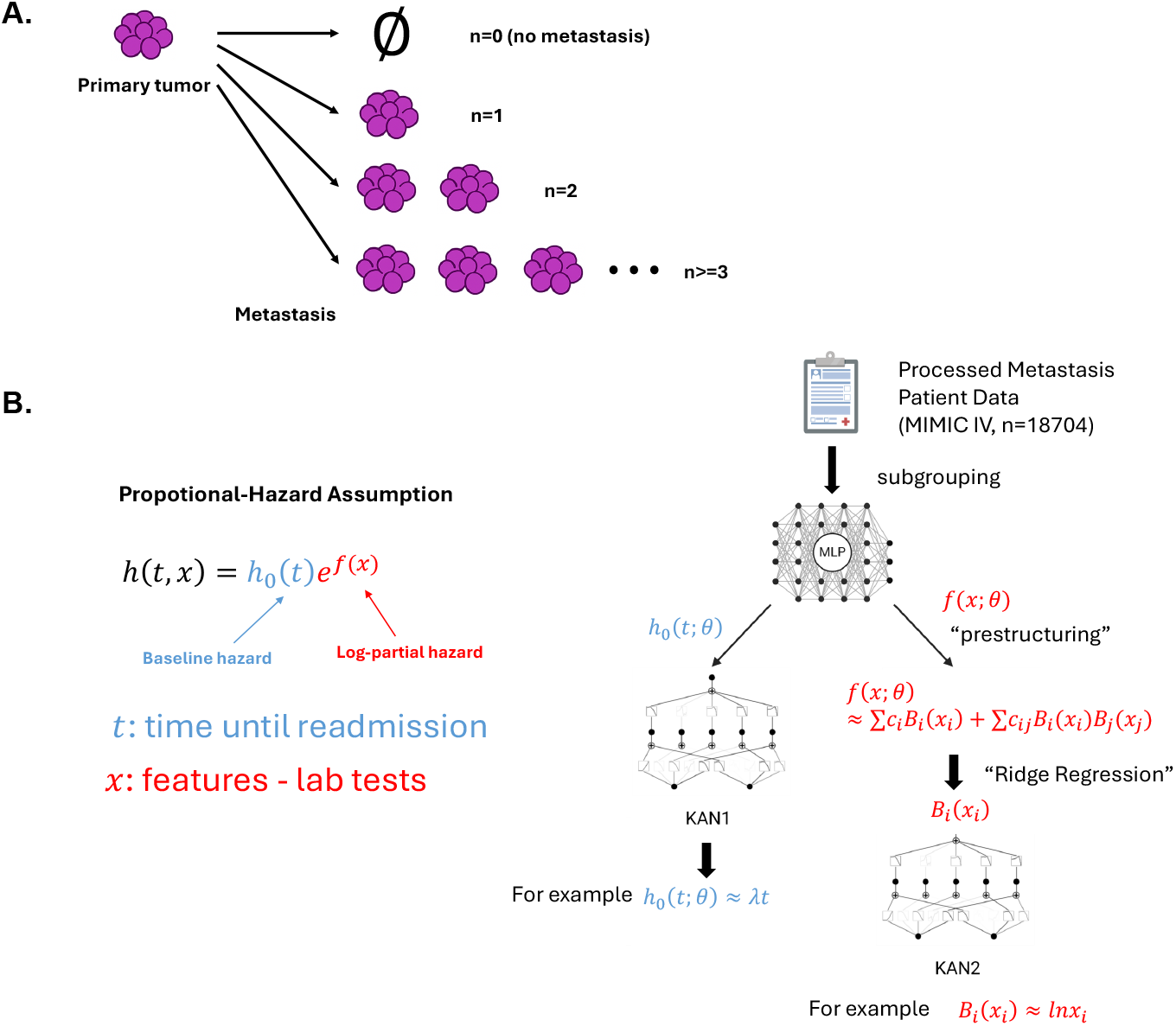
Illustration of metastasis type classification and design of work flow. **A.** According to the number of metastatic sites/tumors(*n*), patients are classified into 4 different groups: *n* = 0*, n* = 1*, n* = 2*, n >*= 3, to represent different risk levels. **B.** Design of work flow. Preprocessed patient data are subgrouped based on metastastic tumor burdens, and then input into an overparameterized Multi-Layer Perceptron(MLP) network, and loss are designed according to the classic “Proportional-Hazard Assumption”, to learn the relationship between baseline hazard *h*_0_ and time until readmission *t*, and the relationship between log-partial hazard *f* and lab-derived features *x. θ* represents trained parameters from MLP. *h*_0_(*t*; *θ*) will be symbolized directly by a Kolmogorov-Arnold Network(KAN). For *f* (*x*; *θ*), a B-spline expansion considering univariate and interaction terms is introduced as “prestructuring”, akin to “preconditioning” in numerically solving large-scale linear equations. New parameters *c_i_, c_ij_* are introduced by ridge regression, to avoid overfitting. Univariate and interaction terms *B_i_, B_ij_* are symbolized individually by new KANs.

### Survival analysis and the proportional hazard assumption

In classical survival analysis, the key object of interest is the hazard function *h*(*t, x*), which denotes the instantaneous rate of readmission at time *t* given patient features *x*. Under the widely adopted proportional hazard assumption, the hazard decomposes multiplicatively into a baseline hazard *h*_0_(*t*) (shared across patients) and a feature-dependent component:

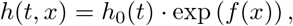

where *f* (*x*) is known as the log-partial hazard. Traditional Cox proportional hazards models assume *f* (*x*) = *β*^⊤^*x*, yielding a linear log-risk in the covariates [16]. The correspondence between model input variables *x_i_* and their associated lab-derived clinical features is summarized in Table 1.

**Table 1.** Mapping between standardized model variables *x_i_* and their corresponding lab-derived clinical features.

| Variable | Lab-Derived Feature Name |
| --- | --- |
| $x_0$ | first_blood_pressure |
| $x_1$ | first_height |
| $x_2$ | platelet_count |
| $x_3$ | white_blood_cell_count |
| $x_4$ | hemoglobin |
| $x_5$ | glucose |
| $x_6$ | bicarbonate |
| $x_7$ | creatinine |
| $x_8$ | chloride |
| $x_9$ | potassium |
| $x_{10}$ | sodium |

### Neural survival model training

To capture nonlinear feature effects and richer survival dynamics, we train a multi-layer perceptron (MLP) to simultaneously learn both the log-partial hazard function *f* (*x*; *θ*) and the baseline hazard function *h*_0_(*t*; *θ*). Following the DeepSurv framework [23, 31], the model is optimized by minimizing the negative hazard-based log-likelihood, averaged over the training set:

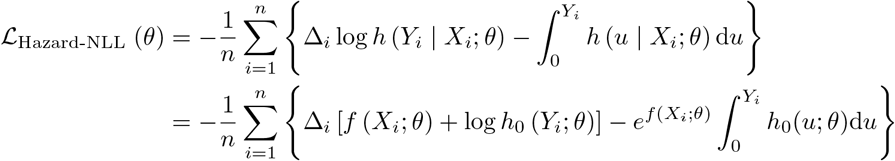

where *Y_i_* is the observed time (i.e., time to readmission), *X_i_* denotes the lab-derived feature vector, and Δ*_i_* = 1 indicates an uncensored observation, while Δ*_i_* = 0 denotes right-censoring. The training objective is to find the optimal network parameters,

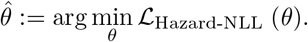

Although the MLP provides a powerful and flexible model, it introduces substantial risk of overfitting and remains largely uninterpretable. These limitations motivate the need for a symbolic distillation phase to extract intelligible and clinically meaningful insights.

### Symbolic distillation with B-spline prestructuring

To extract interpretable formulas from the trained MLP outputs, we first apply a B-spline basis expansion to the lab-derived features. For each feature *x_i_*, we compute a set of *k* B-spline basis functions 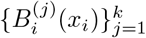 of degree *d*, typically cubic (*d* = 3). We then construct both univariate and pairwise interaction terms:

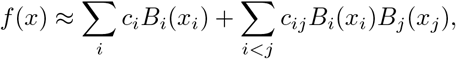

where *c_i_* and *c_ij_* are learned via ridge regression, effectively “prestructuring” the feature space before symbolic extraction.

### Symbolization via Kolmogorov

**Arnold Networks** Finally, to obtain compact symbolic approximations of the log-partial hazard *f* (*x*), we apply

### Kolmogorov

**Arnold Networks (KAN)** and their extension **MultiKAN** [32, 33] to the outputs of the B-spline ridge regression. These symbolic models help distill the high-dimensional interaction-expanded representation of *f* (*x*) into intelligible expressions. In contrast, for the baseline hazard *h*_0_(*t*), which is a univariate function of time, we directly apply KAN without the B-spline expansion. This distinction reflects the different nature of the two components: while *f* (*x*) depends on multivariate lab-derived features requiring structured interaction modeling, *h*_0_(*t*) captures temporal effects that can be learned directly with univariate symbolic approximators. KANs approximate multivariate functions through compositions of additive and univariate nonlinear transformations, and their general structure can be expressed as:

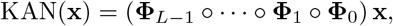

where each **Φ***_𝓁_* denotes a KAN layer, and *L* is the total number of layers. In this formulation, each layer applies learned univariate nonlinear functions to its inputs and aggregates them through summation nodes.

MultKAN extends this architecture by incorporating both summation and multiplication nodes, allowing it to model interactions between input variables directly. This enriched representational capacity makes MultKAN especially suitable for capturing interaction effects that are prevalent in our survival modeling context. Both KAN and MultKAN yield interpretable, closed-form symbolic expressions, making them ideal for distilling the outputs of overparameterized deep models into intelligible and clinically meaningful forms.

### Connection to Cox models

Our final distilled forms can be seen as interpretable generalizations of Cox models. In the special case where *f* (*x*) is linear and *h*_0_(*t*) is unspecified, our model reduces to the classical Cox model. However, our symbolic approach allows recovery of expressive and metastasis-specific hazard forms, enabling nuanced insight into readmission dynamics across subgroups.

## Results

### Initial Analysis

#### Correlation between readmission time, metastastic burden, and lab-derived features

To better understand the relationship between clinical features and cancer readmission, we examined the correlation matrix between `y readmission day`, the number of metastatic tumors, and a set of lab-derived features(See Figure 2). In the correlation matrix, only `chloride`and `sodium`has a relatively high correlation,and that’s even no more than 0.5, and all the other linera corrleations are weak. The observed weak linear correlations between readmission and most individual lab features suggest that univariate or additive linear models are insufficient to capture the underlying predictive structure. Despite the presence of biologically relevant variables (e.g., metastasis count), the lack of strong pairwise associations indicates that the predictive signal may lie in higher-order, nonlinear, or interaction effects. While correlation analysis is a standard preprocessing tool used to detect and eliminate highly redundant features, it is not well-suited for identifying predictive variables, especially in complex biomedical datasets. Features that show weak linear correlations with the target may still carry strong predictive signals through nonlinear transformations or interactions. Our observed weak correlations between lab features and readmission suggest that linear additive models would likely fail to capture the true structure of the data. This supports our use of symbolic regression and spline-based expansions, which are capable of uncovering interpretable nonlinear and interaction effects that traditional correlation-based filtering would overlook.

**Figure 2.**
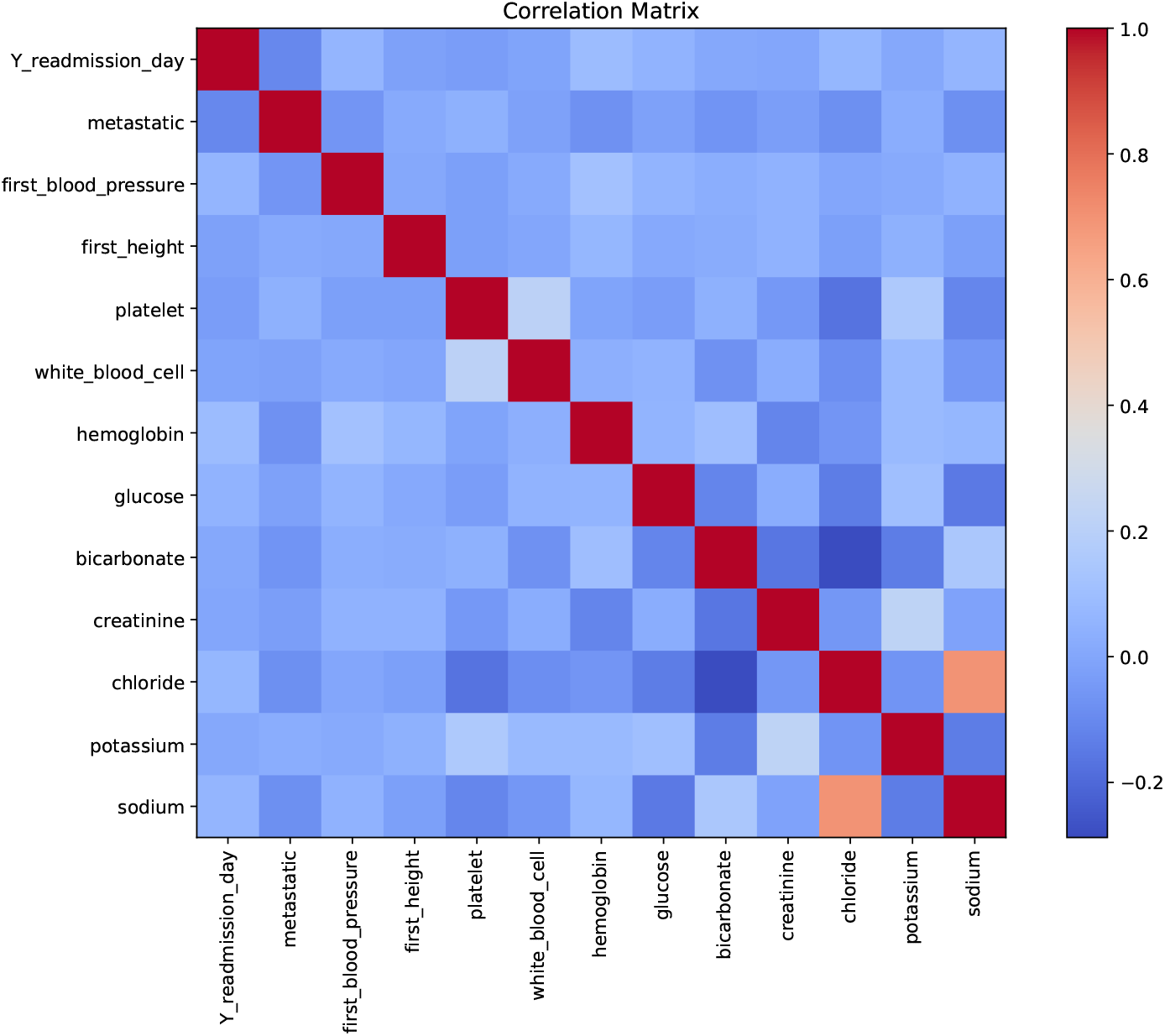
Correlation matrix between readmission time, metastasis burden, and key laboratory measurements. This figure presents the Pearson correlation matrix among the time-to-readmission variable (`Y readmission day`), metastasis count (metastatic), and a panel of clinical features including vital signs and laboratory test results such as blood pressure, platelet count, white blood cell count, hemoglobin, glucose, bicarbonate, creatinine, chloride, potassium, and sodium. While most lab features exhibit modest correlations with each other, the correlation between `Y readmission day`and both metastasis and other lab features appears weak (near 0), suggesting limited linear association.

#### Kaplan-Meier Estimation

To gain an initial understanding of how varying metastatic burdens influence hospital readmission risk in cancer patients, we employed the Kaplan–Meier estimator to visualize *non-readmission probabilities* stratified by the number of metastatic sites (see Figure 3). This non-parametric survival analysis method estimates the probability of remaining readmission-free over time and is defined as:

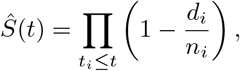

**Figure 3.**
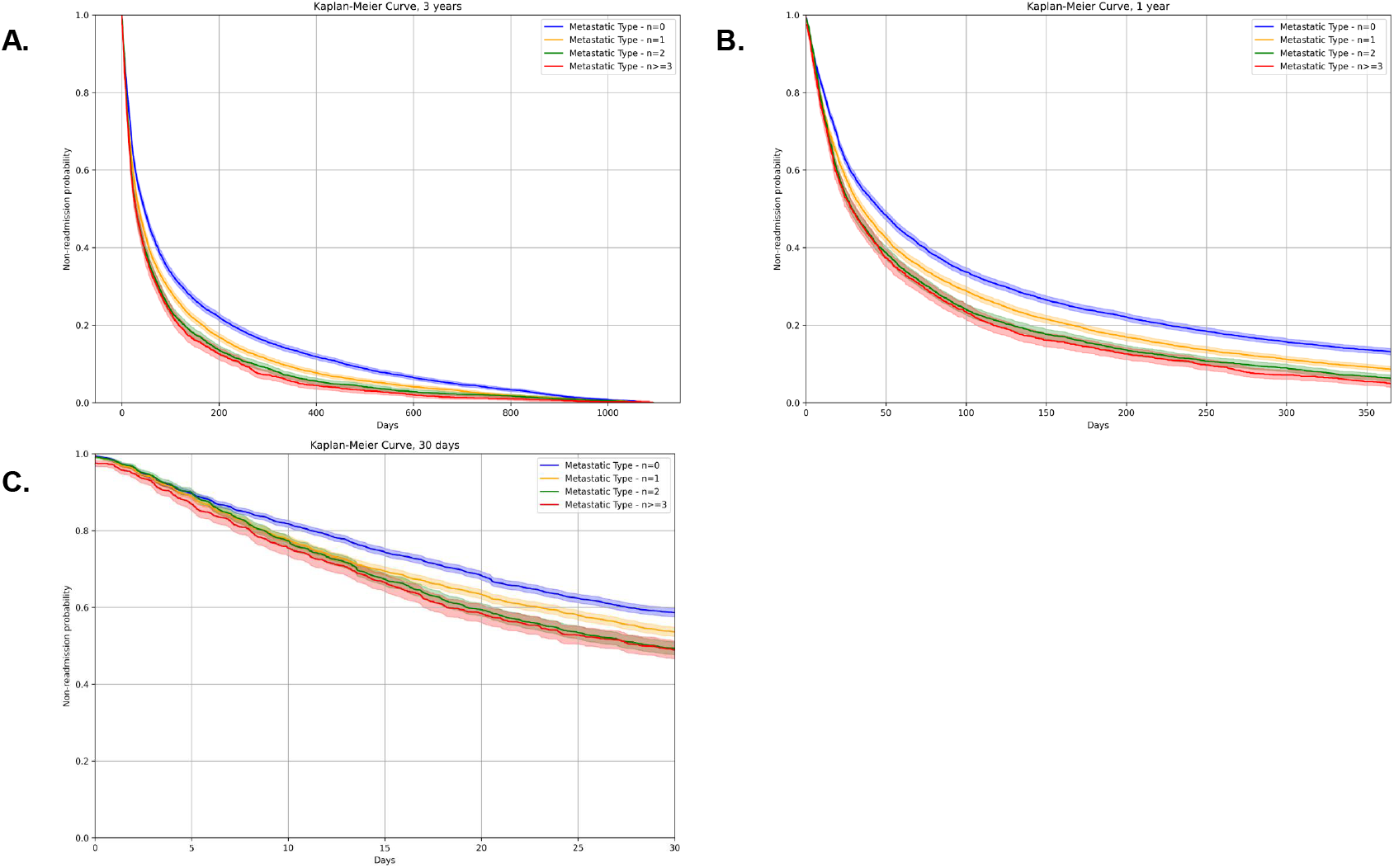
Kaplan-Meier curves of non-readmission probability for 3 years, 1 year, 30 days. **A.** Kaplan-Meier survival curves are plotted to illustrate the probability of non-readmission over time for 4 different patient groups according to metastastic tumor numbers for a total time length of 3 years. **B.** and **C.** is zoomed in from A for a one-year readmission risk and 30-day readmission risk.

where *t_i_* represents the time of a readmission event, *d_i_* is the number of readmissions occurring at time *t_i_*, and *n_i_* is the number of patients at risk immediately prior to *t_i_*. The Y-axis in Figure 3 denotes the estimated probability of not being readmitted after discharge, while the X-axis tracks the number of days post-discharge. In the full 3-year analysis window (Figure 3A), we observe a consistent separation of the survival curves by metastatic burden. Patients without metastases (blue line) exhibit the highest non-readmission probabilities over time, reflecting the lowest risk of early readmission. Those with one metastatic site (orange line) maintain slightly lower probabilities, followed by patients with two (green line) and three or more metastatic sites (red line), both of which exhibit similarly low non-readmission probabilities, nearly overlapping across the full time span. This trend suggests a nonlinear and threshold-like effect of metastatic burden: while the presence of metastases significantly increases readmission risk, the incremental difference between two and three or more metastases is marginal. A closer look at the first year post-discharge (Figure 3B) reveals a steep decline in non-readmission probabilities for all groups. By the end of the first year, patients with any metastases show a near-zero chance of remaining readmission-free. Even for those without metastasis, the non-readmission probability drops below 20%, indicating that most cancer patients are at high risk of readmission within 12 months—especially those with metastatic progression. These findings emphasize the need for targeted post-discharge monitoring and early intervention strategies, particularly within the first year for patients with any metastatic burden. Zooming further into the first 30 days after discharge (Figure 3C)—a critical window for hospital performance metrics and reimbursement penalties—we observe that patients with two or more metastases (green and red lines) follow nearly indistinguishable non-readmission trajectories. This suggests that, from a short-term policy perspective, differentiating between two and three or more metastatic sites may not yield additional benefit in predicting near-term readmission risk. In contrast, patients without metastases remain comparatively more likely to avoid readmission during this initial period.

Taken together, these Kaplan–Meier analyses highlight the profound impact of metastatic burden on hospital readmission likelihood across multiple time horizons. These results support differentiated monitoring strategies: while 30-day readmission risk may not strongly discriminate between different metastatic burdens, the first year after discharge represents a critical window where metastasis-stratified post-discharge planning and interventions may substantially improve outcomes.

### Interpretable Relationships Between Hazard and Lab Tests Inform Readmission Risk in Metastatic Cancer Patients

#### Baseline Hazard Estimation and Interpretation

In this section, we present the training results obtained from the workflow illustrated in Figure 1. After training a multi-layer perceptron (MLP) on the entire dataset for 100 epochs, we achieved consistently high C-index values across all subgroups stratified by metastatic burden, with each subgroup scoring above 0.8 (See Table 2). Given the proportional hazards assumption, which decomposes the hazard function into a time-dependent baseline hazard *h*_0_(*t*) and a covariate-dependent log-partial hazard *f* (*x*), we independently examine and symbolize these two components. It is important to note that our MLP is overparameterized, which may lead to overfitting. Thus, we employ symbolic distillation as a form of regularization to make the learned relationships more interpretable and intelligible [34].

**Table 2.** C-index comparison across methods and metastasis subgroups. Models vary in overfitting risk and interpretability. B-spline expansions include univariate (Univ.) and interaction (Inter.) terms.

| Method | Overfitting Risk | Interpretability | $n = 0$ | $n = 1$ | $n = 2$ | $n \geq 3$ |
| --- | --- | --- | --- | --- | --- | --- |
| 1. Cox Regression | Low | Most interpretable | 0.555 | 0.537 | 0.547 | 0.534 |
| 2. MLP | High | Least interpretable | 0.8969 | 0.9379 | 0.9524 | 0.9648 |
| 3. MultKAN | Low | Most interpretable | 0.5002 | 0.4972 | 0.5036 | 0.4836 |
| 4. Univ. B-spline (Linear) | Low | Interpretable | 0.5632 | 0.5810 | 0.5785 | 0.6248 |
| 5. Univ. B-spline (Linear, same amp.) | High | Interpretable | 0.6734 | 0.6877 | 0.7522 | 0.8718 |
| 6. Univ.+Inter. B-spline (Linear) | High | Interpretable | 0.8198 | 0.8425 | 0.9497 | 0.9648 |
| 7. Univ.+Inter. B-spline (Ridge) | Low | Interpretable | 0.6996 | 0.7150 | 0.7767 | 0.8775 |

Focusing first on the baseline hazard, we examine the one-year post-discharge time frame, as suggested by the Kaplan–Meier (KM) curves in Figure 3, which highlight this period as critical for differentiating post-discharge policies among cancer patients with varying levels of metastasis. In Figure 4, we visualize the numerically estimated *h*_0_(*t*) curves for each metastatic subgroup (*n* = 0, 1, 2, ≥3), along with their corresponding symbolic approximations via KAN. These symbolic equations achieve low training and testing losses on the order of 10^−2^ to 10^−3^. Patients without metastasis (*n* = 0, blue line) exhibit a flat, stable hazard, while those with *n* = 1 or *n* = 2 show similarly low trends with slightly more curvature—represented by quadratic symbolic forms. In contrast, patients with a high metastatic burden (*n* ≥ 3, red line) display a significantly elevated baseline hazard, peaking sharply early on and remaining high throughout the year.

**Figure 4.**
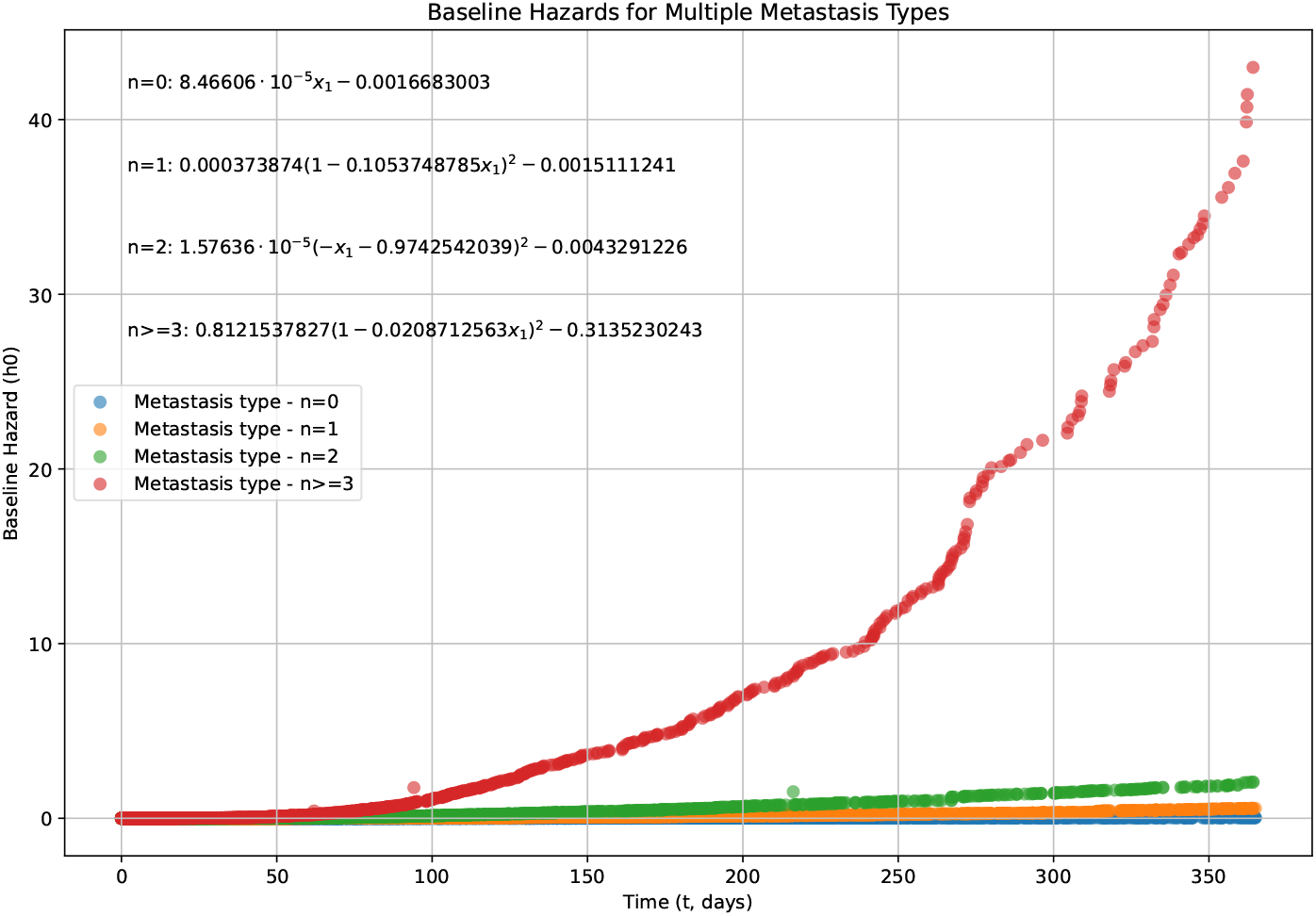
Baseline hazard functions *h*_0_(*t*) estimated for patients with different numbers of metastatic tumors. Each curve represents a metastasis group. *n* = 0, *n* = 1, *n* = 2**, and** *n >*= 3**, showing how baseline risk of readmission evolves over time under each condition.** This figure illustrates the estimated symbolic baseline hazard functions *h*_0_(*t*) over time (in days) for patient subgroups stratified by metastasis count. The hazard functions were learned independently for each subgroup, capturing the temporal dynamics of readmission risk in the absence of patient-specific covariates (i.e., mathematically equivalent to setting *x* = 0 after standardization during data preprocessing). Symbolized function *h*_0_(*t*) are listed on the left top corner.

This divergence between *n* ≤2 and *n* ≥ 3 is notable because, in the KM plots (Figure 3), especially in the 30-day window, the survival curves for *n* = 2 and *n* ≥ 3 appear nearly identical. The apparent discrepancy arises from the different natures of these measures: the KM curve reflects overall cumulative hazard over time, while *h*_0_(*t*) only captures baseline instantaneous risk. One plausible explanation is that although patients with *n* ≥ 3 exhibit high baseline hazard rates, their realized total hazard *h*(*t x*) = *h*_0_(*t*) exp(*f* (*x*)) may be moderated by low-risk feature profiles captured by *f* (*x*). In other words, for many patients in the *n* ≥ 3 group, favorable lab-derived features may suppress the actual readmission risk early on, producing KM curves similar to those with *n* = 2.

This suggests that while elevated baseline hazard is indicative of a generally high-risk subgroup requiring clinical attention, it is not sufficient for determining individual outcomes. Instead, the feature-dependent component *f* (*x*) plays a crucial role in modulating this risk—especially among patients with high metastatic burden(*n >*= 3). For these patients, personalized risk assessments that incorporate both baseline hazard and patient-specific features are essential for guiding post-discharge monitoring and interventions in a one-year window.

#### Comparison Across Modeling Strategies for Log-partial Hazard

As demonstrated in our baseline hazard analysis, it is critical to derive interpretable relationships between the log partial hazard and lab-derived features to enable personalized readmission risk estimation for cancer patients.

To this end, we implemented a B-spline basis expansion with interaction terms, followed by ridge regression, prior to symbolic modeling using KAN or MultKAN. This B-spline prestructuring serves as a powerful initialization strategy for symbolic learning: by projecting input features into a high-dimensional space of univariate and pairwise interaction terms, it transforms nonlinear dependencies into a structured, approximately linear representation. The ridge regression step regularizes this expansion, isolating informative terms while controlling overfitting. This process effectively acts as a preconditioner, reducing the complexity of the optimization landscape and easing the symbolic discovery of concise, intelligible formulas. Rather than learning both structure and coefficients from scratch, symbolic models are guided by an interpretable scaffold that improves convergence and enhances generalization. This framework allows us to decouple complexity from unintelligibility—delivering both high accuracy and symbolic interpretability.

To evaluate the performance and generalizability of our method, we compare it with several baseline models that vary in overfitting risk, interpretability, and predictive accuracy across metastasis subgroups (see Table 2). In addition to our final framework (Method 7), we include six control strategies:

- **Method 1**: Classic Cox proportional hazards regression without deep learning or feature expansion.
- **Method 2**: Plain MLP trained on raw features, without symbolic distillation.
- **Method 3**: MLP followed by MultKAN for direct symbolic learning of *f* (*x*).
- **Method 4**: MLP followed by univariate-only B-spline expansion and linear regression.
- **Method 5**: As in Method 4, but with the same number and scale of B-spline parameters as used in our interaction-based method, to match expressiveness.
- **Method 6**: Full univariate and interaction B-spline expansion fitted with linear regression (without regularization).

We observe that while Method 2 (MLP alone) achieves the highest C-indices, it suffers from significant overfitting and lacks interpretability. In contrast, Method 1 (Cox) and Method 4 (univariate linear regression) are interpretable and low-risk but perform poorly, suggesting that purely additive or univariate models (e.g., generalized additive models [35]) cannot capture the complex interactions underlying readmission risks.

Interestingly, Method 3, which applies MultKAN directly to the MLP, achieves high interpretability but low C-index. This suggests that the loss landscape is too complex for direct symbolic learning without prior structural guidance. Method 6, which fits interaction terms via linear regression, achieves high C-indices but with large coefficients and signs of overfitting. Finally, Method 7, our proposed method incorporating both interaction B-spline expansion and ridge regularization, strikes a favorable balance—achieving high C-indices across all subgroups while maintaining low overfitting risk and interpretability. To ensure that the good performance of Method 7 is not merely a consequence of overparameterization, we introduce Method 5 as a control: a univariate-only B-spline expansion with the same order of parameter amplitude, achieved by increasing the number of knots. By comparing the C-indices of Method 5, Method 6 and Method 7, we observe that Method 5 still falls short in predictive performance, highlighting the necessity of including interaction terms in the model.

These findings underscore the value of combining prestructuring with symbolic distillation. Our framework offers a tractable and interpretable alternative to black-box models, enabling both reliable predictions and deeper clinical insight into how metastatic burden and lab test interactions drive readmission risk.

#### Contributions of Univariate and Interaction Terms

To examine the relative contributions of individual features versus their pairwise combinations in predicting readmission risk, we compared the magnitudes of coefficients derived from B-spline basis expansions across four metastasis subgroups (Figure 5). This comparison helps evaluate whether interactions between features carry more predictive signal than univariate effects alone. Figure 5A displays the absolute coefficient values for each term in the linear ridge regression model following B-spline expansion. The y-axis reflects the magnitude of coefficients—used here as a proxy for feature importance—while the x-axis ranks features within each subgroup. Panel A focuses on univariate terms, representing isolated effects of individual lab test features. In contrast, Panel B shows the coefficients of interaction terms, which capture combined effects between pairs of features. Across all four metastasis subgroups (*n* = 0, *n* = 1, *n* = 2, and *n ≥* 3), the interaction terms dominate: their absolute coefficient values are consistently larger than those of univariate terms. To make this more explicit, Figure 5B quantifies the percentage contribution of each term type. In every subgroup, interaction terms account for approximately 98–99% of the total coefficient magnitude, underscoring their overwhelming importance in predicting readmission risk. These results suggest that in the context of cancer patient readmission, it is not just the level of individual biomarkers that matters, but how they interact with one another. This highlights the clinical value of incorporating pairwise relationships into risk models, especially when dealing with complex physiological systems and heterogeneous patient populations.

**Figure 5.**
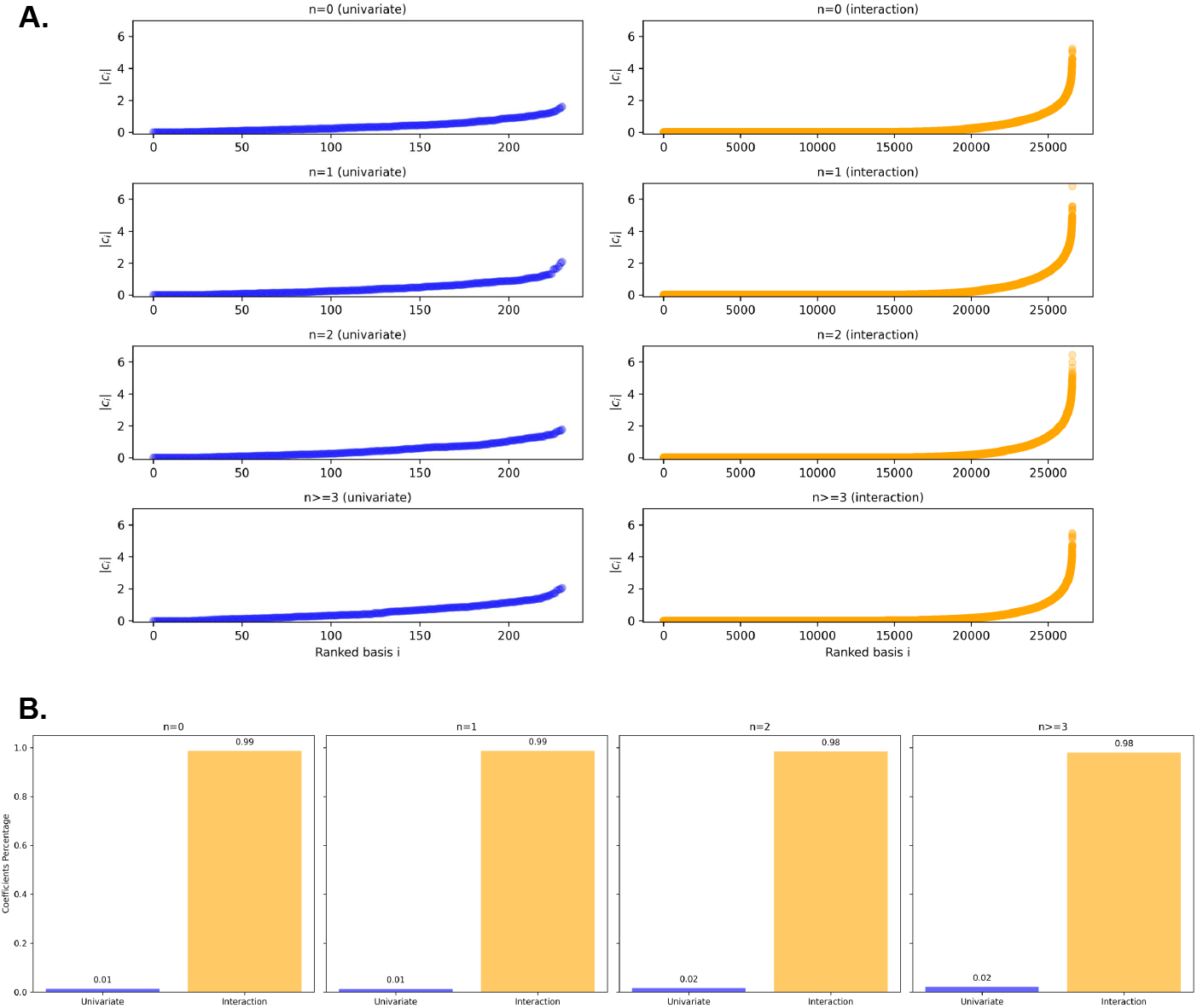
Contribution of Univariate and Interaction Terms Across Cancer Types. **A.** Ranked absolute coefficients of univariate and interaction B-spline terms for four different metastasis subgroups. Each subplot shows a separate metastasis subgroup. Coefficient percentage comparison between univariate and interaction terms, for four different metastasis subgroups.

#### Symbolized Top Terms

By obtaining coefficients from B-spline expansions with interaction terms, we can in theory visualize any relevant feature relationships and distill them using a compact symbolic model such as KAN. Here, we highlight the top 5 terms with the largest absolute coefficients for each metastasis subgroup (*n* = 0, *n* = 1, *n* = 2, and *n ≥* 3) by KAN or MultKAN, as shown in Figure 6. Each symbolic term (KAN(*x_i_*)≈ *B_i_*(*x_i_*), or KAN(*x_i_, x_j_*) ≈ *B_i_*(*x_i_*)≈*B_j_*(*x_j_*)), represents a simplified, interpretable expression derived from B-spline transformed lab test variables—capturing univariate effects or pairwise interactions that contribute most to readmission risk. The visualized outputs (see Figure 6) suggest several important findings. First, across all subgroups, the most predictive terms are interaction terms rather than univariate ones, underscoring the dominance of combinatorial feature effects over isolated predictors in shaping readmission risk. Second, the specific symbolic forms and response surfaces vary significantly across metastasis levels, suggesting metastasis-specific rewiring of interaction structure and risk attribution. This heterogeneity highlights the utility of symbolic learning not only for transparency but also for hypothesis generation—revealing potentially meaningful lab variable interactions modulated by metastatic burden. Third, for each metastasis burden group, `x 0`(corresponding to `first blood pressure`, see Table 1) is the only feature that consistently appears in at least one interaction term. This suggests that while first blood pressure alone may not serve as a strong standalone predictor of readmission risk in cancer patients, its interaction with other routine lab features may carry meaningful prognostic value. Therefore, greater clinical attention may be warranted to blood pressure in conjunction with other biomarkers during discharge planning. For full symbolic expressions of each term, see the Supplementary File `symbolic top terms.txt`.

**Figure 6.**
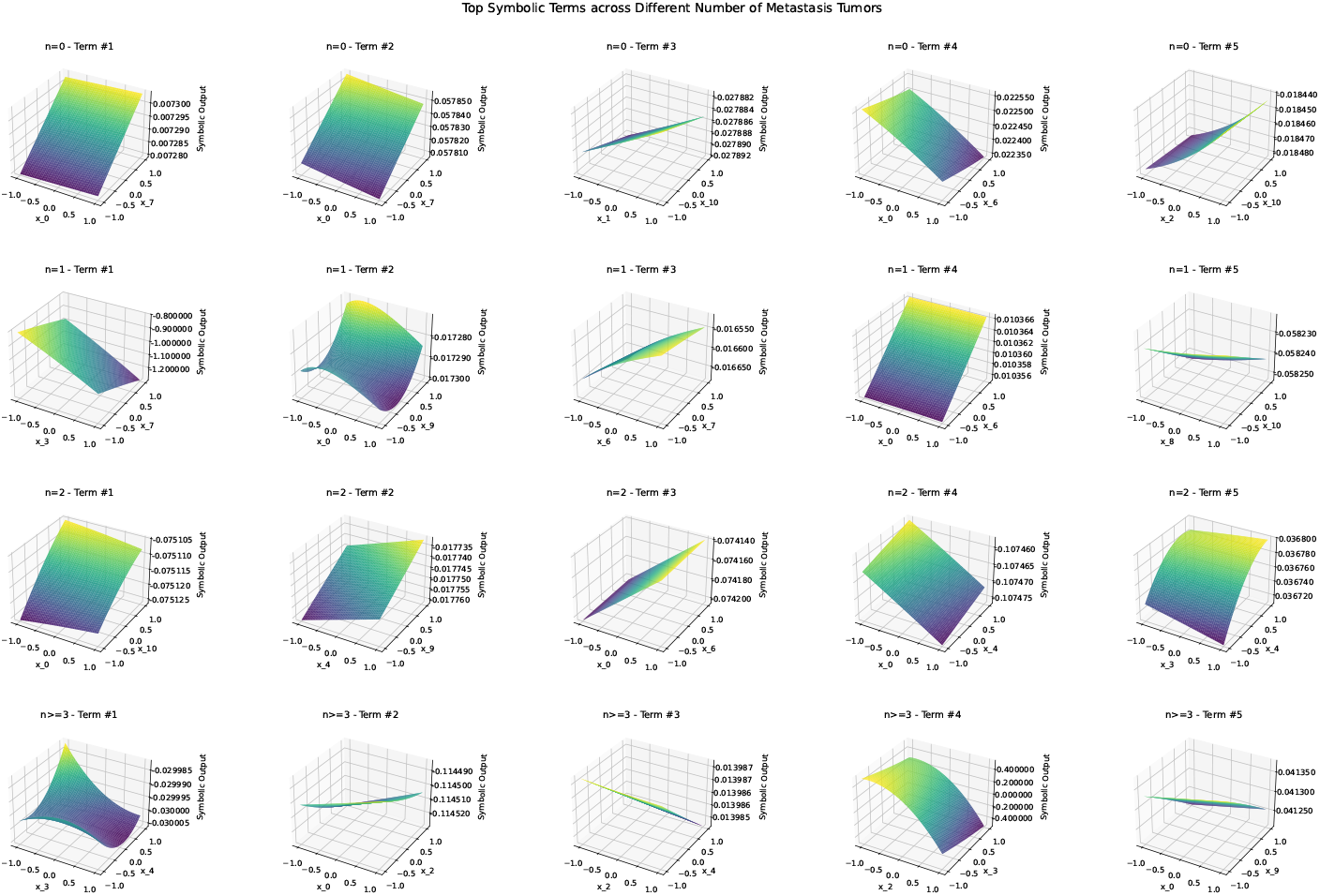
Symbolic representations of top predictive terms for different metastatic burdens. Top five symbolic terms identified from Multi-KAN symbolic distillation for each metastatic category (*n* = 0, 1, 2*, >*= 3). Each subplot visualizes a symbolic function involving univariate or bivariate B-spline basis inputs selected as most predictive under ridge regression and symbolic extraction. The *x*-and *y*-axes represent standardized lab-derived features, while the *z*-axis corresponds to the symbolic output value of the interaction term. Note that the output is shown without applying the corresponding regression coefficient weight.

## Discussion

### Summary of Our Contribution

In this study, we proposed a fast, interpretable, and symbolic framework for predicting readmission risk among cancer patients with varying metastatic burdens by combining a multi-layer perceptron (MLP), Kolmogorov–Arnold Networks (KAN), and a B-spline expansion with interaction terms. Our method decouples the hazard function into baseline and covariate-dependent components, and then distills each into simplified symbolic expressions. Despite minimal hyperparameter tuning and high levels of clinical noise, the framework achieved a competitive C-index, demonstrating its ability to uncover both predictive and clinically plausible patterns.

These findings offer actionable insights for hospital post-discharge strategies:

- **30-day Readmission:** Readmission risk within 30 days does not strongly discriminate between patients with different metastatic burdens. Therefore, a uniform post-discharge policy in this early window may be cost-effective while maintaining adequate care.
- **One-year Window:** The first year after discharge represents a critical period where stratified intervention based on metastatic burden may be more impactful. Specifically, patients with higher metastatic burden (e.g., ≥3 metastatic sites) exhibit significantly elevated baseline hazard. However, our model also shows that their realized risk is shaped by interactions among lab-derived features. Hence, hospital resources should be directed toward personalized monitoring and care, prioritizing high metastatic-burden patients identified as high-risk through such feature interactions.

### Advantages of Our Framework

Our framework requires no heavy hyperparameter tuning or deep model regularization. Yet, even in this undertuned setting, it successfully identified interpretable symbolic relationships that generalize across subgroups and align with clinical intuition. Recent models such as CoxKAN [36] utilize one-step Kolmogorov–Arnold Networks (KANs) with multiple regularization terms to generate symbolic representations of the log-partial hazard function. However, these outputs remain complex and often challenging to interpret. Furthermore, due to the current implementation limitations described in their paper, CoxKAN does not incorporate MultKAN and therefore can only capture univariate effects. This restricts its expressive power by excluding multiplicative interactions between features, effectively reducing it to a form of Generalized Additive Models (GAMs) [35] or Neural Additive Models (NAMs) [37], where the hazard is modeled as *g*(𝔼[*f*]) = *β* +∑ *_i_ ϕ_i_*(*x_i_*). In contrast, our proposed framework explicitly models both univariate and pairwise interaction effects through a two-step symbolic distillation approach. By first applying B-spline expansion and ridge regression as a prestructuring layer, and then distilling with Kolmogorov–Arnold Networks, we enable interpretable symbolic expressions that include feature interactions. This approach not only improves modeling flexibility but also offers a lightweight, modular alternative for hypothesis generation and exploratory analysis in noisy clinical datasets. We also observed clear differences in both baseline and log-partial hazard functions across metastatic burdens. Patients with three or more metastatic sites consistently exhibited steeper hazard trajectories. These results suggest that standard laboratory tests can reflect distinct survival dynamics when interpreted in the context of metastasis burden.

### Scientific Implications: The Role of Interactions

Our symbolic expressions highlight that readmission risk cannot be captured by any single laboratory test alone. Instead, nonlinear combinations—especially pairwise interactions of B-spline components—are key drivers of predictive accuracy. Notably, the importance of these pairs shifts across metastasis strata, suggesting a modulating role of metastasis. There is empirical support from additive models. Our findings align with prior work on Generalized Additive Models with Pairwise Interactions (GA^2^M), in the form of 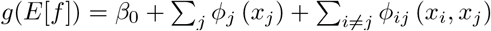
[38], which have been shown to match black-box models in clinical tasks like ICU mortality, pneumonia risk, and hospital readmission prediction while retaining transparency [34]. In particular, Caruana et al. demonstrate that GA^2^M recovers meaningful feature combinations that clinicians can interpret and trust. Actually, olw-order structure is sufficient and natural. The success of additive + interaction models may reflect a general property of natural systems, where second-order effects dominate and higher-order interactions are often unnecessary [39]. Similar principles have been observed in neuroscience, genetics, and biophysics [40–43], and now appear relevant to cancer readmission risk modeling.

### Limitations and Caveats

First, in terms of dataset size and external validity, our dataset is modest and lacks external validation. As such, the exact symbolic expressions derived from our models should not be overinterpreted. We emphasize that the broader structural patterns—such as which lab features consistently appear in high-impact interactions—are likely to be more robust than individual coefficients or symbolic terms. Second, the study is limited by missing or aggregated clinical variables. Several well-established prognostic biomarkers based on lab-derived feature combinations—such as the Neutrophil-to-Lymphocyte Ratio (NLR) and the Pan-Immune-Inflammation Value (PIV)—have clearly defined symbolic equations (see Supplementary Information). However, due to the absence of key components (e.g., neutrophil or monocyte counts), these scores could not be directly calculated from our dataset. Even if such features were available, our relatively simple symbolic learning framework may not be expressive enough to recover these complex clinical indices. Nonetheless, these classic biomarkers could serve as useful benchmarks or constraints in future symbolization efforts. Lastly, our current model does not account for cancer subtype heterogeneity. Patients were grouped solely by metastatic burden, without stratification by cancer origin or site-specific metastasis. Future work could incorporate cancer subtype information to improve specificity and uncover more granular interaction patterns between lab-derived features and disease progression.

### Conclusion and Clinical Implications

Despite these limitations, our framework yields several robust and clinically actionable insights: 1. Patients with ≥ 3 metastatic sites are at significantly elevated readmission risk and should receive targeted post-discharge monitoring. 2. Pairwise combinations of lab features, not individual tests, better capture metastasis-specific readmission dynamics. 3. Symbolic expressions vary across metastasis burden, indicating distinct biological and clinical pathways. We recommend that hospital readmission risk models consider metastasis-aware stratification and explicitly incorporate interaction effects between laboratory features. Our symbolic framework may also serve as a foundation for developing better interpretable, lab-based scoring systems in oncology care.

## Data Availability

All data produced in the present study are available upon reasonable request to the authors.

## Acknowledgments

N/A.

## Supplementary Information

### Training Details

#### MLP Structure

We implement a multilayer perceptron (MLP) comprising an input layer of size 11 (corresponding to the 11 lab-derived features), followed by five hidden layers with 1024, 1024, 1024, 512, and 512 neurons, respectively. Each hidden layer uses ReLU activation, and the final output layer has a single neuron with no activation. The architecture is summarized as:

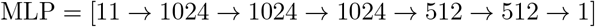

Bias terms are included in all layers except the final output layer. The total number of trainable parameters is 3,016,456. Given the dataset size of 18,704 uncensored admissions, the MLP is clearly overparameterized, underscoring the importance of symbolic distillation for interpretability and generalization.

#### B-spline Structure

- To model nonlinear relationships among lab features, we employ a cubic B-spline basis expansion (degree = 3) with 20 uniformly spaced internal knots for each univariate feature. This yields a smooth and flexible representation of the input space without requiring prior knowledge of functional forms.
- To capture second-order feature interactions, we generate pairwise interaction terms by taking the elementwise product of B-spline basis functions from unique feature pairs. This structured approach enables modeling of nonlinear feature interactions.
- The univariate basis expansion produces (20 + 3 + 1) × 11 = 24 × 11 = (264) features. The number of unique feature pairs among *d* = 11 features is 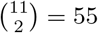 Each pair produces 24 × 24 = 576 interaction terms, resulting in a total of 55× 576 = 31, 680 interaction features. In total, the B-spline expansion with both univariate and interaction terms yields 264 + 31, 680 = 31, 944 features.
- For comparison, a univariate-only B-spline model with an equivalent parameter count was constructed using 2000 internal knots (for a total of (2000 + 3 + 1)× 11 = 2004 × 11 = 22, 044 parameters), thereby matching the representational capacity of the interaction-expanded version.
- To mitigate overfitting due to high dimensionality, Ridge regression is applied to the expanded feature matrix to regularize the model coefficients prior to symbolic regression.

#### KAN Structure

- For univariate functions such as the baseline hazard *h*_0_(*t*) or single-feature transformations *B_i_*(*x_i_*), we use a single-layer Kolmogorov–Arnold Network (KAN) with one neuron. The grid and polynomial degree are set to `grid=3`, `k=3`, and random seed `seed=42`. Optimization is performed using the `LBFGS`algorithm with `steps=50`and `lambda=0.001`. Symbolic expressions are rounded using the `ex round`rule to 10 decimal places.
- For modeling interactions between feature pairs (*x_i_, x_j_*), we apply the MultiKAN framework, which extends KAN by including multiplicative neurons. The network consists of an input layer with two neurons, a hidden layer with five standard neurons and one multiplicative neuron, and an output layer. The configuration is summarized as width=[2,[5,1],1], with the same grid and polynomial settings as above. The base activation function is set to `identity`, and multiplicative arity is set to 2.
- Across symbolic training tasks for both univariate and interaction terms, the training and testing losses consistently converge to values on the order of 10^−2^ or lower, indicating good approximation quality with compact symbolic expressions.

## Metrics

### Concordance Index (C-index)

Model performance was assessed using the concordance index (C-index), a standard metric in survival analysis that quantifies the agreement between predicted risks and actual event times [31]. The C-index ranges from 0.5 (random guessing) to 1.0 (perfect prediction). For each metastatic burden subgroup and modeling approach described in the main text, we computed the corresponding C-index. Mathematically, the C-index is defined as:

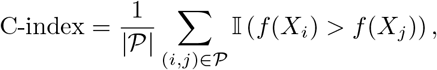

where𝒫 = {(*i, j*)| *Y_i_* < *Y_j_,* Δ*_i_* = 1} is the set of comparable pairs (i.e., where subject *i* experienced an event before subject *j* and is uncensored), *f* (*X*) denotes the predicted risk score (corresponding to the log-partial hazard in our framework), and 𝕀(·) is the indicator function that evaluates to 1 if its argument is true and 0 otherwise.

### Symbolic Training and Testing Loss in KAN

To evaluate the quality of symbolic approximations obtained via KAN or MultKAN, we compute the symbolic loss, which reflects the mean squared error (MSE) between the predicted symbolic output *y_i_* and the original target value *y_i_* from the overparameterized MLP or ridge regression model. Both training and testing symbolic losses are reported. Formally, the symbolic loss is given by:

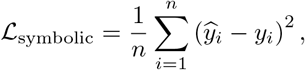

where 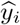 is the output of the learned symbolic expression (e.g., from KAN), and *y_i_* is the ground-truth numerical function value (either *h*_0_(*t*), or *B_i_*(*x_i_*)). In our experiments, symbolic losses are typically on the order of 10^−2^, indicating a close approximation.

### Common Cancer Prognostic Scores and Analytical Equations

This section summarizes widely used prognostic scores in oncology, grouped by their input features [44–62]. These scores help assess systemic inflammation, nutritional status and liver function.

#### A. Inflammation-Immune Indices

- **1 Pan-Immune-Inflammation Value (PIV) [44–46]***Purpose:* Composite immune marker reflecting systemic inflammation.

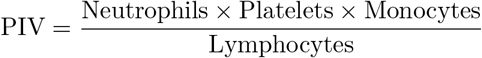
- **2 Neutrophil-to-Lymphocyte Ratio (NLR) [47, 48]***Purpose:* Indicator of pro-vs. anti-inflammatory balance.

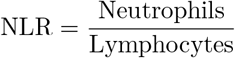
- **3 Platelet-to-Lymphocyte Ratio (PLR) [49, 50]***Purpose:* Marker of platelet activation and immune suppression.

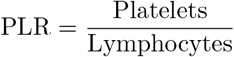
- **4 C-Reactive Protein to Albumin Ratio (CAR) [51, 52]***Purpose:* Quantifies inflammation relative to nutritional status.

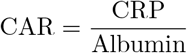

#### B.Nutritional and Functional Status Scores

- **5 Prognostic Nutritional Index (PNI) [53, 54]***Purpose:* Reflects nutritional and immune competence.

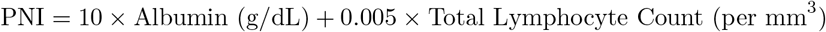
- **6 Body Mass Index (BMI) [55, 56]***Purpose:* Overall nutritional status and obesity classification.

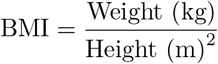

#### C. Liver and Metabolic Function

- **7 Glasgow Prognostic Score (GPS) [57, 58]***Purpose:* Evaluates inflammation and hepatic protein synthesis.
  ‐ CRP larger than 10 mg/L and Albumin less than 3.5 g/dL: Score = 2
  ‐ CRP larger than 10 mg/L or Albumin less than 3.5 g/dL: Score = 1
  ‐ Otherwise: Score = 0
- **8 Child-Pugh Score [59, 60]***Purpose:* Assesses liver function severity using 5 clinical measures: Albumin, Bilirubin, INR, Ascites, Encephalopathy. *Scoring:* Each variable is scored from 1 to 3; total score determines class A–C.
- **9 Anion Gap [61, 62]***Purpose:* Evaluates acid-base balance.

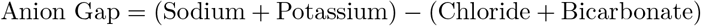

## Summary

Many of these prognostic scores are constructed through specific feature combinations or interactions, reflecting domain knowledge distilled into interpretable formulas. As such, they provide valuable benchmarks for evaluating the effectiveness of symbolic learning approaches in predicting readmission risk. Future research could explore whether such empirically derived symbolic scores can be rediscovered or approximated through data-driven learning, thereby bridging clinical intuition and machine-derived insights.

## Supplementary Files

The symbolic expressions corresponding to the top five terms—ranked by the absolute magnitude of their coefficients—for each metastasis subgroup are provided in the supplementary file `symbolic top terms.txt`.

